# Severe airport sanitarian control could slow down the spreading of COVID-19 pandemics in Brazil

**DOI:** 10.1101/2020.03.26.20044370

**Authors:** Sérvio Pontes Ribeiro, Wesley Dáttilo, Alcides Castro e Silva, Alexandre Barbosa Reis, Aristóteles Góes-Neto, Luiz Carlos Junior Alcantara, Marta Giovanetti, Wendel Coura-Vital, Geraldo Wilson Fernandes, Vasco Ariston C. Azevedo

## Abstract

**Background:** We investigated a likely scenario of COVID-19 spreading in Brazil through the complex airport network of the country, for the 90 days after the first national occurrence of the disease. After the confirmation of the first imported cases, the lack of a proper airport entrance control resulted in the infection spreading in a manner directly proportional to the amount of flights reaching each city, following first occurrence of the virus coming from abroad.

**Methodology:** We developed a SIR (Susceptible-Infected-Recovered) model divided in a metapopulation structure, where cities with airports were demes connected by the number of flights. Subsequently, we further explored the role of Manaus airport for a rapid entrance of the pandemic into indigenous territories situated in remote places of the Amazon region.

**Results:** The expansion of the SARS-CoV-2 virus between cities was fast, directly proportional to the airport closeness centrality within the Brazilian air transportation network. There was a clear pattern in the expansion of the pandemic, with a stiff exponential expansion of cases for all cities. The more an airport showed closeness centrality, the greater was its vulnerability to SARS-CoV-2.

**Conclusions:** We discussed the weak pandemic control performance of Brazil in comparison with other tropical, developing countries, namely India and Nigeria. Finally, we proposed measures for containing virus spreading taking into consideration the scenario of high poverty.

## Introduction

In the last few weeks, the new disease COVID-19 has been spreading rapidly around the world mainly due to stealth transmission, which started in China at the end of 2019. Large continental countries are likely to be very vulnerable to the occurrence of pandemics (Morse et al. 2012). While the dissemination dynamics have varied between regions, country sanitary policies play a key role in it. For instance, two very large developing countries, India and Brazil, have a very different epidemical pattern. On March 18^th^, India had 137 cases and Brazil 621, as recorded in the Brazilian Ministry of Health and John Hopkins monitoring sites dedicated to SARS-CoV-2 and COVID-19. From 17^th^ to 18^th^ March, Brazil had an increase of 31% in one day, with only four capitals exhibiting community transmission, which was the same to India. However, a very distinct pattern in the ascending starting point for the reported disease exponential curve was observed in each country. By enlarging the comparison to another developing tropical country in the Southern Hemisphere (thus in the same season), we selected Nigeria, since it was the first country to detect a COVID-19 case in Africa. Nigeria displayed less than 10 confirmed cases during the same period of time. Furthermore, Nigeria has a population (206 million) similar to that of Brazil (209 million).

Both India and Nigeria claim they imposed severe entrance control, and close following up of each confirmed case, as well as their living and working area, and people in contact with them. In Brazil, the Ministry of Health has developed a good monitoring network and a comprehensive preparation of the health system for the worst-case scenario. Nonetheless, apparently, the decisions from the Ministry of Health did not cover airport control, and only on March 19^th^, eventually too late, the government decided to control the airports, avoiding the entrance of people coming from Europe or Asia. Hence, the entrance of diseased people in Brazil has been occurring with no control, at least until the aforementioned date. Moreover, after confirming that a person is infected with SARS-CoV-2, his/her monitoring is initiated but there is no monitoring of his/her living network.

For pandemic situations, such as that with which we are living with SARS-CoV-2, the classical algebraic ecological models of species population growth from Verhulst, and species interaction models from Lotka-Volterra, are theoretical frameworks capable to describe the phenomenon and to propose actions to stop it (Pianka 2000). In many aspects social isolation is a way to severely reduce carrying capacity, i.e., the resources available for the virus dissemination. This is the best action for within-city pandemic spreading of coronavirus (Hellewell et al. 2020), since the main form of transmission is direct contact between people or by contact with fomite, mainly in closed environments, such as classrooms, offices, etc. (Rothe et al., 2019; Bedford et al., 2020). Regardless of virulence, for a highly contagious virus such as SARS-CoV-2, the occurrence of the first case in a nation will result in a strongly and nearly uncontrollable exponential growth curve, depending only on the number of encounters between infected and susceptible people, and fuelled by a high H0 (the number of people one infected person will infect).

On the other hand, the dynamics of disease spreading among cities are entirely distinct. In this work, we present an epidemiological model describing the free entrance of people coming from two highly infected countries with close links to Brazil: Italy and Spain. We showed how SARS-CoV-2 spreads into the Brazilian cities by the international airports, and then to other, less internationally connected cities, through the Brazilian airport network. For exploring the dynamics of a continent size, nationwide spreading of SARS-CoV-2, as it is the case of Brazil, we assumed cities connected by airports formed a metapopulation structure.

Each person in a city was taken as a component of a superorganism, i.e., an interdependent entity where living individuals are not biologically independent between them in various subtle ways. By doing so, we dealt with cities as the sampling units, not the people. Flights coming from foreign countries with COVID-19 (namely Spain and Italy for this article) represent the probability of an external invasion of infection in each city. Additionally, we also further explored the vulnerability of the Amazon region, especially of those remote towns where indigenous and traditional communities predominate.

## Materials & Methods

In order to describe the pattern of air transportation and its role in the spreading of the disease, we built a SIR (Susceptible-Infected-Recovered) model (Hethcote 1989; Anderson 1991) split amongst the cities that are interconnected by flights. In this model, the population size inside each city is irrelevant, as well as when the collective infection stage was reached. Thus, we assumed that the city was fully infected and became infectious to the whole system, and, therefore, became a source and not a sink of infection events. Hence, the SIR model started having cities with only susceptible events. Infected events only appeared by migration, i.e. travelers only from Italy and Spain, for sake of simplicity and proximity to the facts.

After the first occurrence is registered in the country, infected events started to spread through the national airlines.

We used a modified version of the SIR model, which took into account the topology of how the cities-demes were linked by domestic flights. In the SIR original model, the infection of susceptible cities occurs by probability *β* of a healthy being (*S*) encounters an infected one (*I*). Conversely, the model has a probability of an infected one get recovered (*R*) given by a parameter *γ*. Analytically:

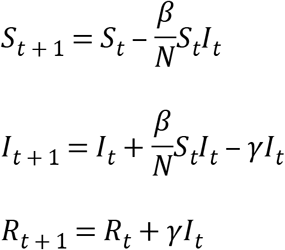

where the indexes *t* and *t*+1 represent the present time and the next time, respectively, and *N*=*S*+*I*+*R* is the total constant population. In this work, we proposed two modifications of the SIR model. The first one is related to the fact that we considered all Brazilian cities that have an airport. Thus, we had *S*^*i*^, *I*^*i*^, and *R*^*i*^ where *i* was a given city. In our case study, 1≤*i*≤154. Another important modification was that related to the connections among airports or cities. Using ANAC data, it was possible to track all the domestic flights in Brazil (Figure 1): https://www.anac.gov.br/assuntos/dados-e-estatisticas/historico-de-voos

**Figure 1.**
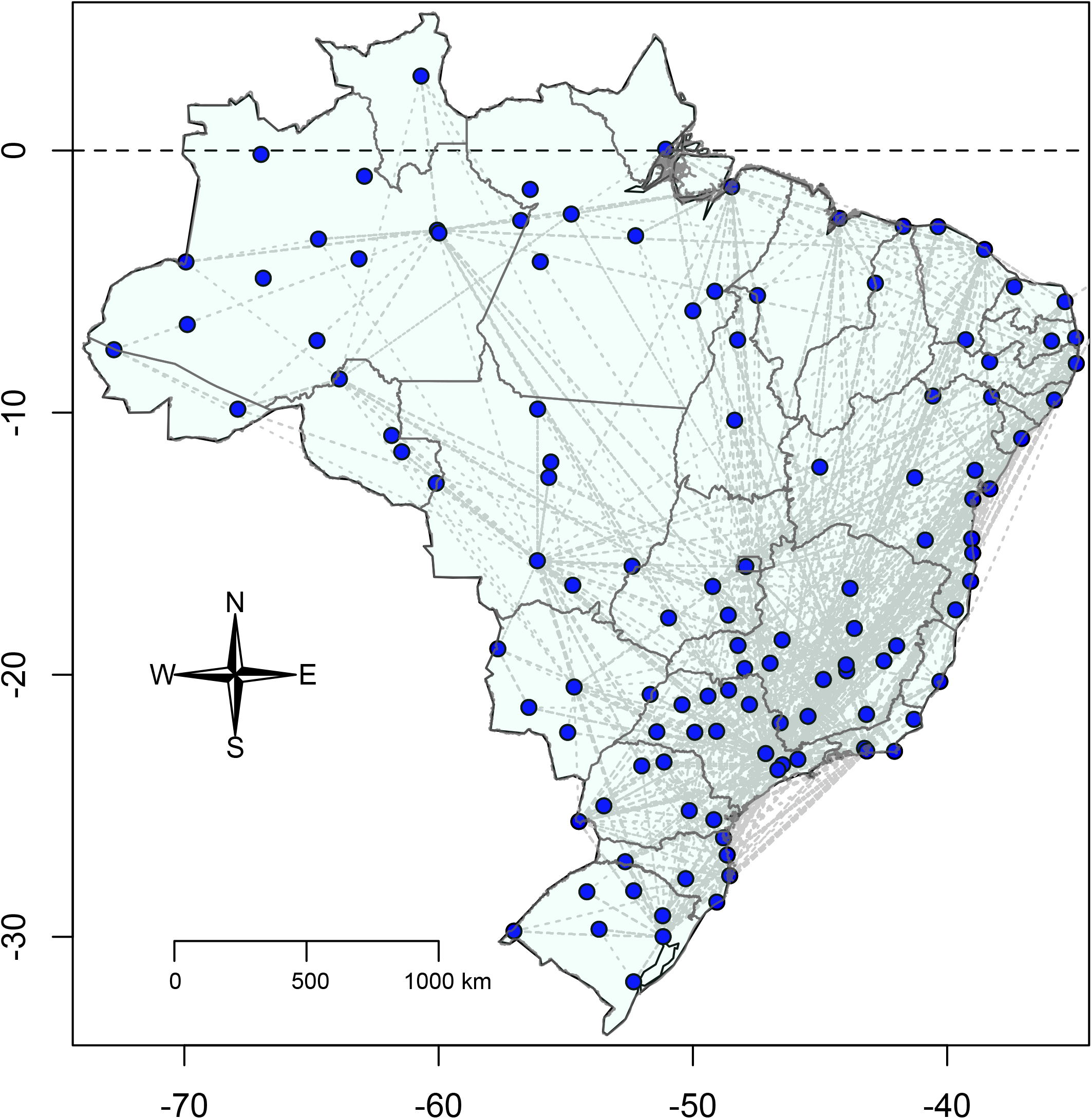
Brazilian flight network, taken from ANAC database.

The modified version of SIR model is then described as follows:

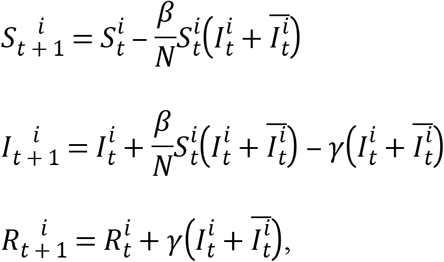

where the upper index *i* indicates the city, and *t* the time. The term 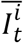 represents the infection added to the *i*^*th*^ city due to traveling diseases, and it is calculated as follow:

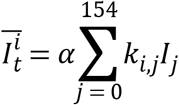

where *k*_*i,j*_ is the number of flights departing at city *i* and arriving at city *j*, and *α* is a newly introduced parameter, which represents the fraction of traveling infected population. For the time, we estimated 90 days for the disease expansion and assumed *γ* as 0, in other words, no recovery. Despite the artificiality of this assumption, we considered that the amount of people still to be infected is larger than those recovered and, thus, becoming resistant, which makes the resistance irrelevant to our output. The model was developed in C and is available as Supplementary Material 1 (and the database as Supplementary Material 2). In addition, we also used a linear model to test whether those cities with higher airport closeness centrality (i.e., important cities for connecting different cities within the Brazilian air transportation network) were more vulnerable to SARS-CoV-2 dissemination.

## Results

The expansion of the SARS-CoV-2 virus between cities was fast, directly proportional to the airport closeness centrality within the Brazilian air transportation network. The disease spread from São Paulo and Rio de Janeiro to the next node-city by the flight network, and in 90 days virtually all the cities with airport(s) were reached, although it occurred with a distinct intensity (Figure 2, Supplementary Material 3). There was a clear pattern in the expansion of the pandemic, with a stiff exponential expansion of cases (measured as the cumulative percentage of infected people per city) for all the cities. On average, the model showed an ascendant curve starting at day 50 (around 15 April), with the most connected cities starting their ascendant curve just after 25 days, and the most isolated ones from day 75 (10th May; Figure 3A). Looking at the daily increment rates, it is clear a first and high peak of infections in the hub cities, happening around 50 days and, starting from 75 days, a new peripheric peak (Figure 3B).

**Figure 2.**
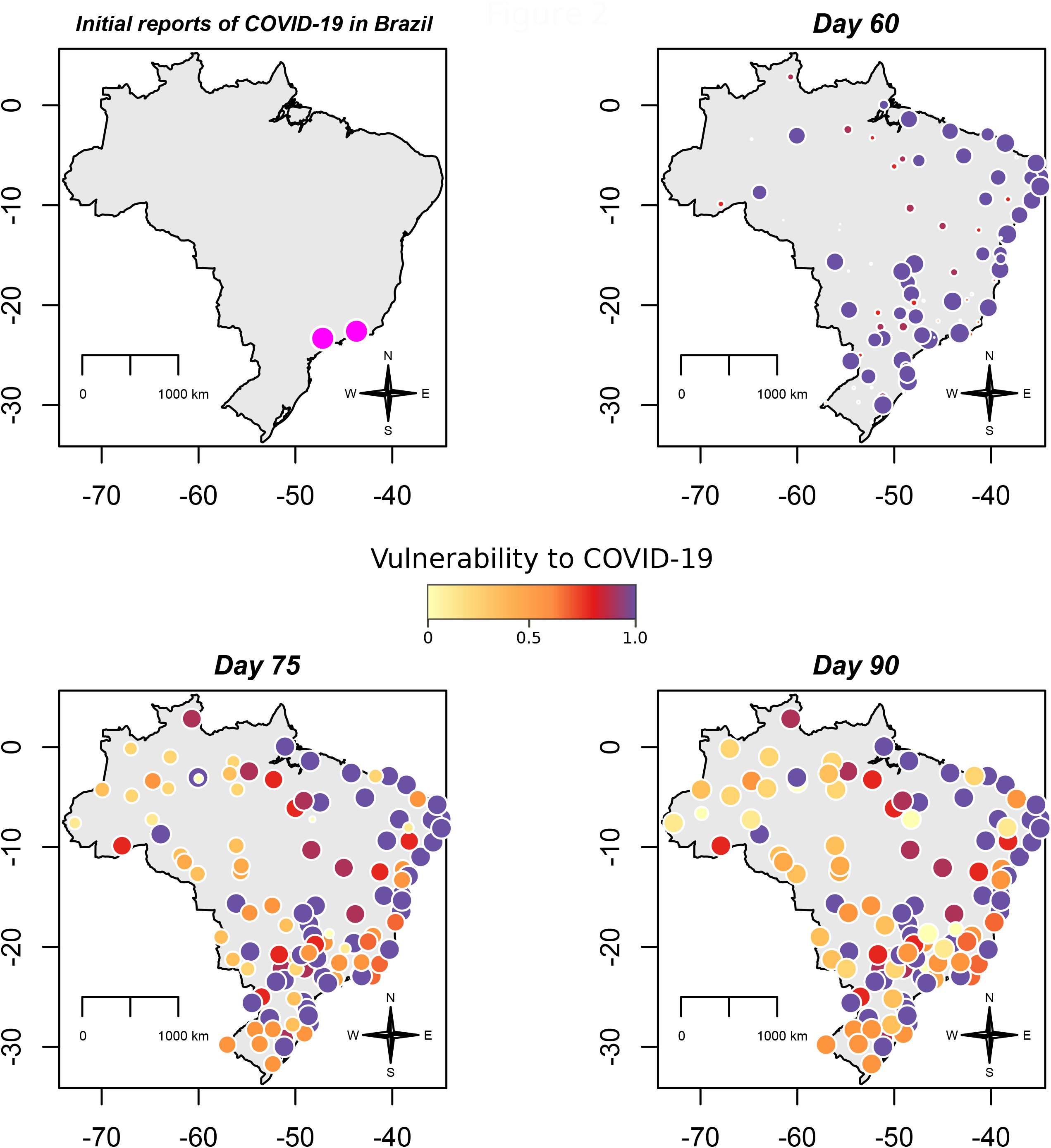
Proportion of infected population of each Brazilian city in 40, 50, 70, and 90 days. Circle colour temperature represents a gradient in percentage of the infected population. Circle size also reflects the size of the pandemics locally in the logarithm scale.

**Figure 3.**
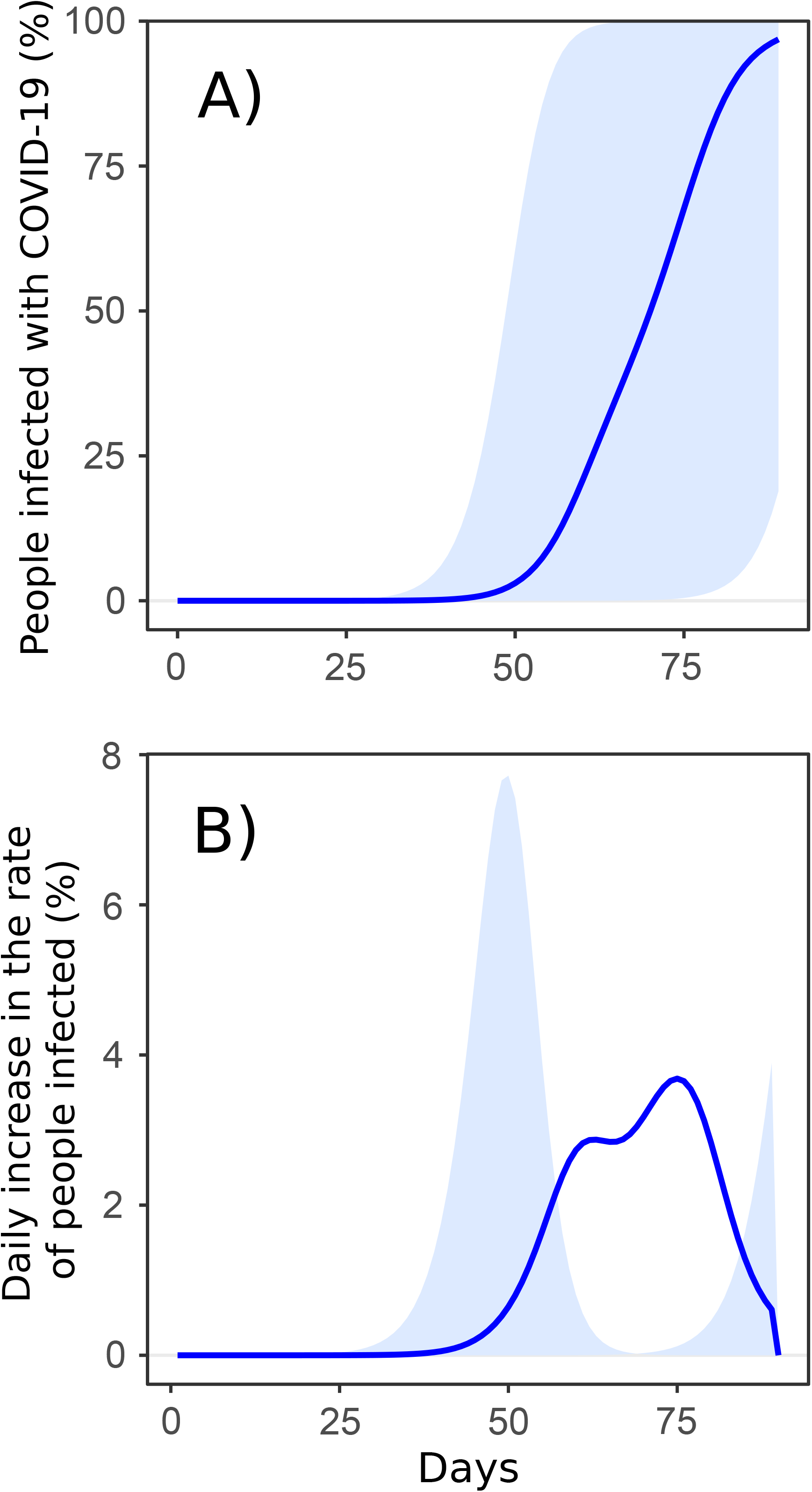
Proportion of infected people per cities until 90 days. A) Cumulative increment rate. The blue line is the national average, and the shadow area is the summing up of minimum and maximum values of all the cities per time interval; B) Daily increment rate. The blue line is the average, showing the overall high rate of infection occurring from 50 to 80 days. Shadow shows the first and the highest peak in the hub cities, around 50 days, and, subsequently, a peripheric peak after 75 days.

The first ten cities to ascend infection rates (São Paulo, Rio de Janeiro, Salvador, Recife, Brasília, Fortaleza, Belo Horizonte, Porto Alegre, Curitiba, and Florianópolis) will actually reach this point about the same time, which is a concerning pattern for the saturation of the public health services. Also, this peak in those cities will saturate all the best hospitals in the country simultaneously.

Therefore, we defined the average proportion of infected people for the 90 days as a measure of vulnerability to COVID-19 dissemination. Henceforth, we found that more an airport shows closeness centrality within the air transportation network, the greater was its vulnerability to disease transmission (Figure 4). This scenario confirmed the importance of a city connecting different cities within the Brazilian air transportation network and, thus, acting as the main driver for the pandemic spreading across the country.

**Figure 4.**
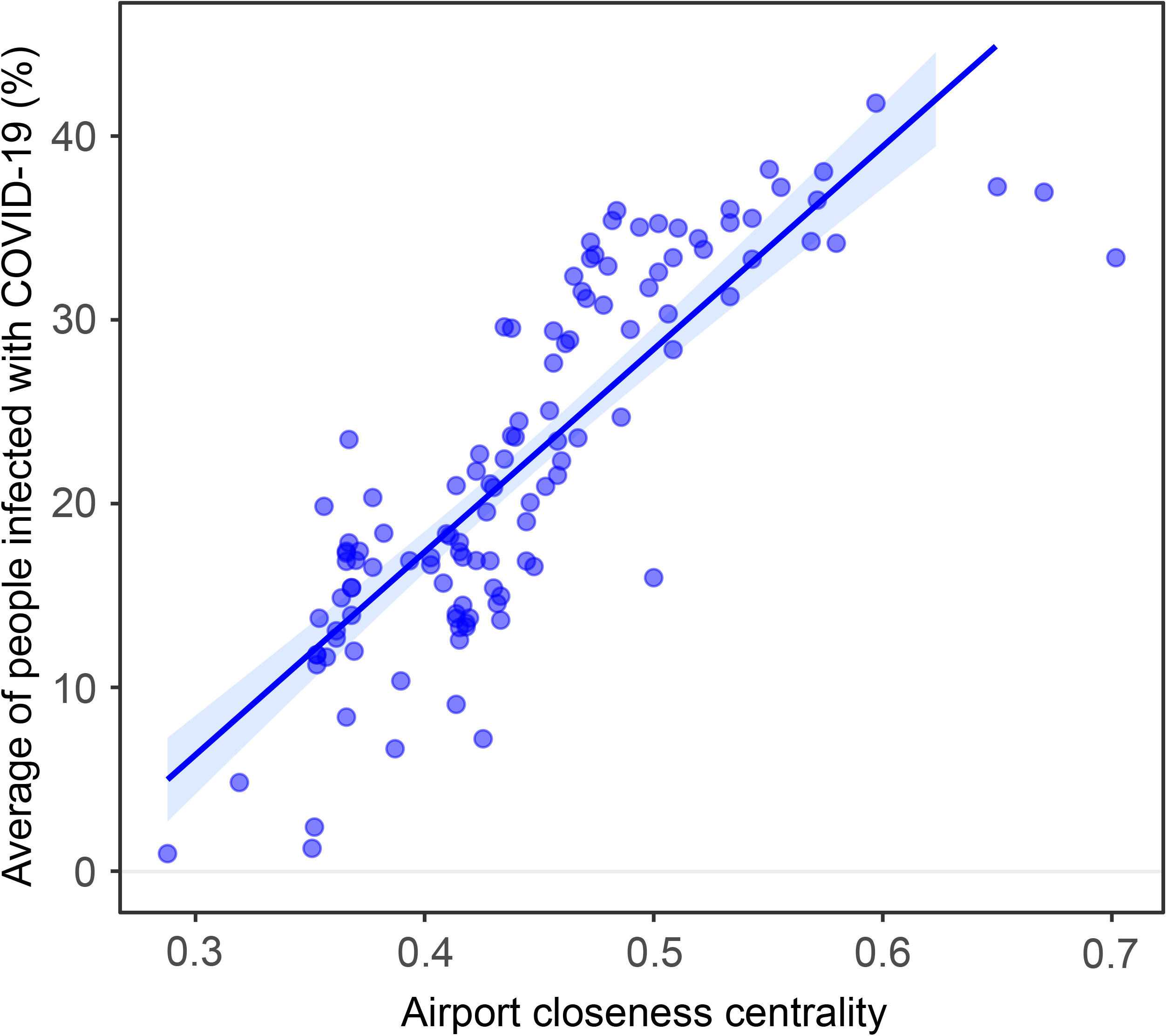
Airport closeness centrality within the Brazilian air transportation network, and its effect on the vulnerability of each city (represented by the average of the percentage of cases per city for the whole 90 days running: r^2^ = 0.71 p < 0.00001).

### Consequences for the Amazonian cities and indigenous people

Herein we showed that an uncontrolled complex airport system made a whole country vulnerable in few weeks, allowing the virus to reach the most distant and remote places, in the most pessimistic scenario. According to our model, any connected city will be infected after three months. As the number of flights arriving in a city is the driver for the proportion of infected people, Manaus, which is a relevant regional clustering, was infected sooner. Indeed, on the 17^th^ of March, Manaus was the first Amazonian city with confirmed cases (without community transmission yet), and it is a node that is one or two steps to all the Amazonian cities. Thus, according to our model, Manaus may reach 1% of the infected population by the 44^th^ day, while, for instance, the far west Amazonian Tabatinga will take 61 days to reach the same 1% of the population infected. By day 60, Manaus may have an average of 50% of its population infected if nothing is be done to prevent it. Tabatinga may also reach the aforementioned value by day 78, if nothing is be done to avoid it. To sum up, within 46 days all the Amazonian cities will have 1% of their population infected and a mean of 50% by day 70.

## Discussion

Brazil has failed to contain COVID-19 in airports and failed to closely monitor those infected people coming from abroad, as well as their living network. One main reason for this is the difficult logistics required to produce such control in a continental country, such as Brazil, which has a complex national flight network. According to the Brazilian Airport Authority, Brazil has the second-largest flight network in the world (just after the USA), with a total of 154 airports registered to commercial flights of which 31 are considered international. In comparison, airport control may be much easier to set up in Nigeria (31 airports of which only five are international). However, with a population 6.4 times higher than Brazil, India, in turn, has a similar sized airport network to Brazil, harboring a total of 123 airports of which 34 are considered international.

Nevertheless, the situation of COVID-19 in India is currently much milder than in Brazil, and it is hard to blame the complexity of the airport networks for the contrasting exponential curve of these two countries. In 20 days from the first infection in Brazil (February 26^th^) against 47 days after the first Indian case (January 30^th^), Brazil already had 5.4 more confirmed cases than India. Clearly one country is doing much better in preventing the entrance of cases and the spreading of the disease by controlling infected citizens. Considering the high probability of a synchronizing SARS-Cov-2 high spreading in various capitals, the country may face a quick health service collapse.

Besides the within-city pattern of virus spreading, one must take into account the pattern of dispersion between cities after the virus has invaded. Additionally, for the Brazilian case, one cannot ignore that, eventually, the occurrence of the first case may have occurred nearly one month before official records, during the carnival period. This is the largest popular street party on the planet, with 6.4 million people in Rio de Janeiro, and 16.3 million in Salvador where the Brazilian Ministry of Tourism revealed that 86,000 foreigners from France, Germany, Spain, Italy, UK, and the US had visited. Considering a disease with so many asymptomatic cases, it could have invaded before but, with the lack of an early warning and airport control, one will never know exactly if this happened. As airport control might have been even more lax in small airports, it might unavoidably result in strengthening of the capability of an infected city to infect the next new one, if no public policy is adopted.

Without a social isolation policy, virus propagation may result in chaotic dynamics, *sensu* May (1976). The lack of control for these situations may result in a dramatic rate of host infection, and an eventual collapse of the host-parasite interaction in a given population, depending on the amount of susceptible, infected and recovered events. Nonetheless, if the population is split into deme-cities, in a metapopulation structure, the collapse takes longer, and a much greater amount of people in different locations may eventually be infected, as found in our model. It is worthwhile to mention that this model, already pessimistic, did not consider the road network, one of the largest on the planet. Most importantly, the best road-connected cities are exactly those mostly connected by airport, and that will be vulnerable earlier, thus, probably spreading the disease faster than our model can predict, unless roads are soon blocked for people. Another weakness of the model is that it cannot quantify a great number of small airports not registered for commercial flights, very common in the Amazonian and Western regions. Taking this into a global scale, for a highly interconnected human population, the consequences may be catastrophic, as it was for the influenza pandemic (Spanish flu) in 1918 (Ferguson et al. 2003). Furthermore, one aspect that must not be neglected is the way as an increasing number of infected people in a city drive the pandemic towards the next city or country. In this context, the complex and large flight network of Brazil, which is also key for the whole Latin America, if not properly monitored and controlled, may cause a window of opportunity for the virus to spread over the entire continent.

The consequences of this uncontrolled SARS-Cov-2 spreading is particularly serious if one takes into consideration the chances of a mutant virulent strain appearing and spreading into poorer and little monitored places of the world. Specifically, for the Amazon region, the lack of any control will make the city of Manaus a very sensitive cluster for public health, due to predominantly poor and indigenous-dominated cities in the region, which are connected to Manaus and will be rapidly infected. Reaching isolated regions means reaching indigenous or traditional communities, whose individuals are classically more susceptible to new pathogens than western-influenced or mixed urban populations. Therefore, a way to prevent such spreading, if still there is time, would be to deal with airports as entrances that need severe infection barriers.

## Conclusions

The eventual lesson to take is that inflexible, severe, and easy to repeat controlling protocols must be applied to all the cities with airports. Likewise, the follow-up monitoring of suspicious individuals and their living network should be reinforced as a national strategy to prevent a large territory to be taken over by a pandemic in a short period of time. In other words, internationally accepted procedures must be taken and even be reviewed to adjust to complex national flight networks of any country. Such procedures must be considered as a priority for national remote airports too, in order to keep poorer and worse equipped cities away from a rapid spread of a pandemic disease.

It is clear at this point that a fast spread of the SARS-CoV-2 is a reality in Brazil, and across most of the country. We proposed this model in order to emphasize the fragility of Brazilian surveillance in the airport network, in an attempt to cause some policy change in time to preserve at least the most remote regions, which are also the most vulnerable, with a weaker health service. Moreover, most of the Eastern part of the country must stay in social isolation in order to prevent a health public collapse by mid-April, as the Ministry of Health predicted. In addition, we also could consider the generalized poverty of Brazil as a further problem our model did not deal with. The chances to produce home-to-home isolation, even legally imposed, is impossible for these poor communities. Nonetheless, considering the few main entrances of most of the Brazilian shanty towns and communities, a similar to airport entrance severe control must be considered to protect a larger but closely connected set of people, eventually following the protocols used for control of Ebola during the last epidemic in Africa (Lau et al. 2017).

## Data Availability

I state all data will be available in submission material

## Acknowledgements

We thank Christina Vinson and Thomas C.A. Williams for the English revision. CNPq agency guarantee research grant scholarship to SPR, ABR, AGN, ACJA, GWF and VAA.

Supplementary Material 1 – Code description - SIR model under network topology. The code was developed in C, and it works as a modification of SIR model running along with the topology of the domestic flight network. After initiating all variables to an initial condition, that is, S (health), I (infected) and R (recovered) of each city, the code starts loading the network and calculates the total number of flights among all the cities. This information is used to feed the classical SIR model introducing in the variable I, the information regarding infected travelers and non-travelers, and the model calculates the next S, I and R of all the cities. This calculation is done in a loop time representing days, the time step that the model was calibrated.

Supplementary Material 2 – ANAC database of aerial transportation network. The spreadsheet presents all the 120 cities with airport(s), their state, latitude and longitude, followed by the closeness centrality in the network. The columns t0 to t90 are the times from 0 to 90 days. Lines for the time columns are the percentage of infected people per city per time.

Supplementary Material 3 – Movie of the spreading of SARS-CoV-2.

This file has a short movie describing the dynamics of SARS-Cov-2 dissemination across Brazil, in two versions.

